# Absence of Care Among Community-Living Older Persons with Dementia and Functional Limitations: A Cross-National Analysis of Population Survey from 22 Countries

**DOI:** 10.1101/2023.10.05.23296622

**Authors:** Zhuoer Lin, Yuting Qian, Thomas M. Gill, Xiaohui Hou, Shanquan Chen, Xi Chen

## Abstract

**Background:** The provision of long-term care for persons living with dementia (PLWD) who have functional limitations is a significant global public health challenge. However, there is limited evidence on the patterns of care received by PLWD across countries and regions. This study aimed to examine the global trends in the absence of care for PLWD with functional limitations and identify potential sociodemographic disparities.

**Methods:** We used harmonized longitudinal survey data from four Health and Retirement Global Family of Studies that surveyed community-living persons aged 50 and older in the United States, England, 18 European countries and Israel, and China. The analysis focused on persons who reported functional limitations and developed dementia during the study periods (2012-2018). Functional limitations were assessed using activities of daily living (ADL) and instrumental activities of daily living (IADL). Absence of care was evaluated as the proportion of PLWD receiving no care for their ADL/IADL limitations.

**Results:** At least 20% of PLWD in both developed and developing countries received no care for their functional limitations, and this absence of care remained stable over time. The absence of care was notable for both ADL and IADL limitations, as well as for informal and formal care. Moreover, substantial disparities were observed, with less-educated individuals and those living alone experiencing greater absence of formal and informal care, respectively. These patterns and trends were consistent across all countries and regions studied.

**Discussion and Implications:** The findings underscore the pressing need to ensure basic care provision for persons with dementia and functional limitations, especially for those who are less educated or living alone. Policymakers should prioritize addressing these disparities and improving care provision for this population worldwide.

## 1. Introduction

The World Health Organization (WHO) estimates that about 16% of the global population experiences disabilities, with a significant portion of them facing functional limitations that impact their day-to-day activities.^1,2^ This prevalence of disability is particularly pronounced among older adults, imposing a substantial demand for caregiving, especially in developing nations.^1,2^ As the population ages and demographics shift, the provision of long-term care for disabled people has become a pressing public health challenge. Notably, the projected quadrupling of older persons who are unable to independently care for themselves by 2050 highlights the potential for major societal impacts.^3^

The burden of disability and the imperative for long-term care are particularly salient for people living with dementia (PLWD), as they often encounter functional limitations as a prominent manifestation of their disabilities.^4,5^ As of 2022, the global count of PLWD exceeded 55 million, with projections indicating an escalation to 139 million by 2050.^6^ Due to the extended duration of illness, PLWD often experience years of disability, spending a substantial portion of their time in a state of severe disability and dependence.^4,5,7^ The financial costs of caring for PLWD worldwide are estimated to exceed 1.3 trillion US dollars annually, and such costs continue to increase.^4,8^ Caregiving needs and healthcare costs among PLWD are also substantially higher than those associated with other conditions such as heart disease and cancer.^9,10^ Moreover, a considerable number of PLWD are older adults dwelling within communities and living alone, which exacerbates the challenge of accessing essential care services.^11,12^

To the best of our knowledge, no studies have evaluated the patterns of care received by PLWD with functional limitations on a global or regional scale across multiple nations.^13^ This lack of information is concerning, as assistance with functional limitations in daily living is crucial for those with dementia. The proportion of PLWD who have functional limitations but do not receive corresponding care is not well understood, and the absence of care may lead to avoidable hospitalization, early institutionalization, heightened mortality risk, increased societal healthcare costs, lowered quality of life.^14–17^ Furthermore, there may be considerable variations over time and across countries in both the need for and the receipt of care among PLWD, making it challenging to generalize existing evidence. Therefore, it is imperative to identify common and differential trends in the absence of care for this especially vulnerable population to inform more effective public policy and interventions.

This study used population-based survey data on community-living adults from the United States (US), England, 18 European countries and Israel, and China to investigate global trends in the proportion of PLWD who receive no care for their functional limitations, as measured by basic or instrumental activities of daily living (ADL/IADL). We hypothesized that 1) a significant proportion of PLWD receive no care for their functional limitations, and the absence of care has changed little over time; 2) PLWD with limited economic resources are more likely to report ADL/IADL limitations without receiving any formal care; 3) PLWD with limited access to care resources (e.g., lower socioeconomic status, living alone) are more likely to report ADL/IADL limitations without receiving any care.

## 2. Methods

### 2.1 Study Design and Participants

We used data from four HRS-family longitudinal surveys, which collected harmonized sociodemographic, economic, health, and cognition data for community-living adults from more than thirty countries. The surveys included the Health and Retirement Study (HRS) in the US;^18^ the English Longitudinal Study of Ageing (ELSA);^19^ the Survey of Health, Ageing and Retirement in Europe (SHARE), which encompasses 28 countries;^20^ and the China Health and Retirement Longitudinal Study (CHARLS).^21^

The four surveys have been designed with similar study protocols and frameworks to facilitate cross-national comparisons, which encompass both developed and developing countries. Specifically, the HRS is a nationally representative longitudinal survey of Americans aged 50 and older, conducted biannually since 1992 with approximately 20,000 respondents per wave.^18,22^ The ELSA includes a nationally representative sample of adults aged 50 and older in England, with biannual rounds since 2002 with around 10,000 participants per wave.^19,23^ SHARE surveys European adults aged 50 and older, starting biannually since 2004, initially covering 10 European countries but subsequently expanding to about 28 countries (including 27 European countries and Israel) in 2017. The SHARE sample size has exceeded 65,000 since 2013.^20^ Lastly, CHARLS is a nationally representative longitudinal study of about 19,000 Chinese adults aged 45 and older, initiated in 2011/2012.^21,24^ In these surveys, each participant completed a standardized questionnaire that was administered face-to-face or via internet/telephone. Further details regarding the sampling and the study protocols can be found in the respective sources.^18–24^

We constructed variables using harmonized HRS-family study data adapted from the RAND HRS and Gateway to Global Aging.^25,26^ These publicly available data sources were harmonized to enhance comparability across the studies. Because only de-identified data were used, our study was exempt from institutional review board review. Participants in the original studies gave informed consent, and each study was approved by a relevant ethics body.^18–24^

To investigate functional limitations and absence of care for older adults with dementia, in each country/region, we limited our samples in each wave to those who reported functional limitations. Moreover, we narrowed down our samples to those who had developed dementia during the study period. To ensure the comparability of measures across surveys, we focused our analysis on data from waves 11-14 (2012-2018) of HRS, waves 6-9 (2012-2018) of ELSA, waves 5-7 (2013-2017) of SHARE, and waves 2-4 (2013-2018) of CHARLS. The final sample range from 2012 to 2018 and included adults aged 50 and over, with 1,229 persons (2,750 person-waves) from the HRS, 493 persons (1,174 person-waves) from the ELSA, 3,377 persons (5,625 person-waves) from the SHARE (covering 19 countries; see Supplementary Table S1), and 1,034 persons (2,071 person-waves) from the CHARLS (Figure 1).

**Figure 1.**
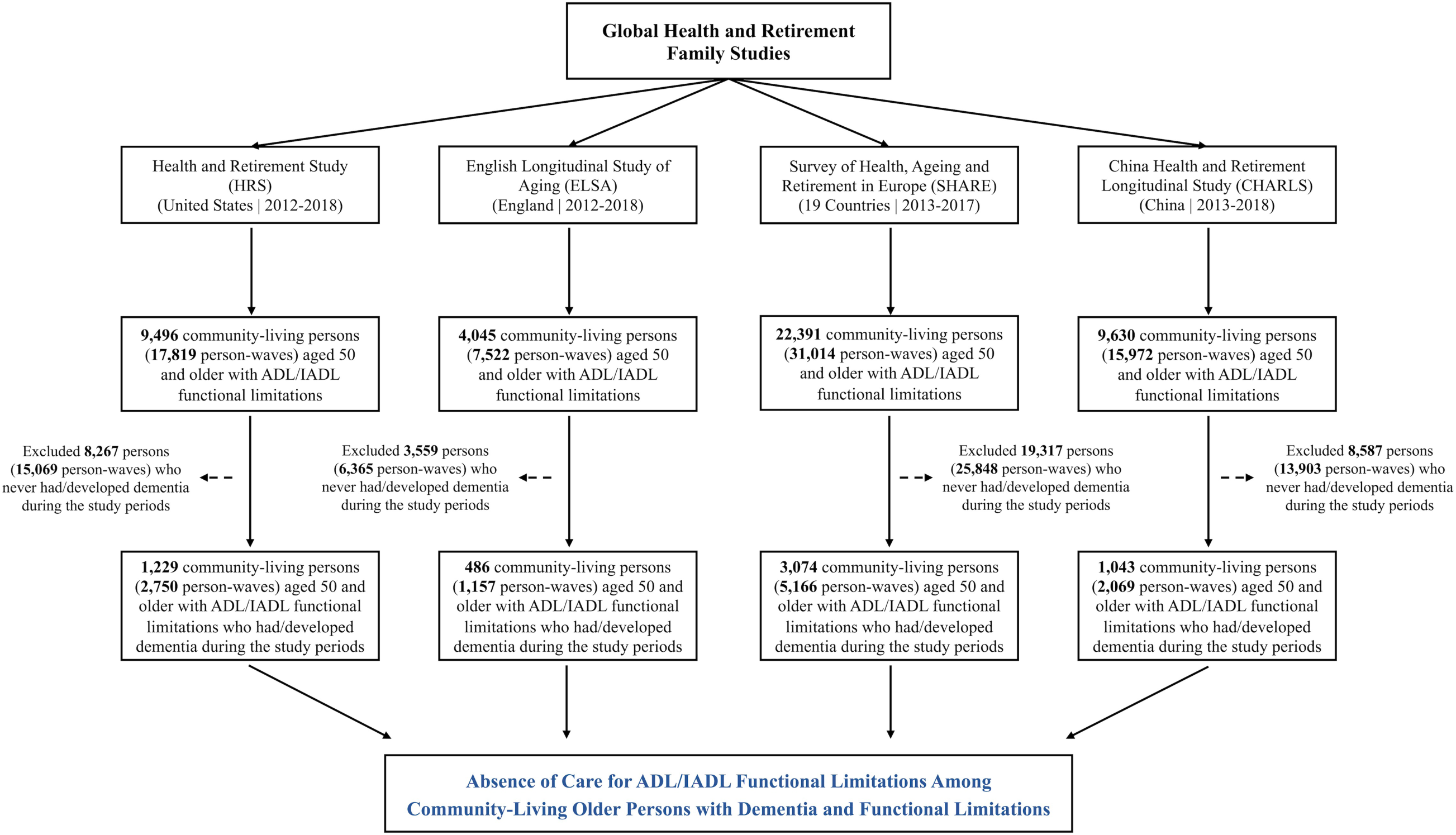
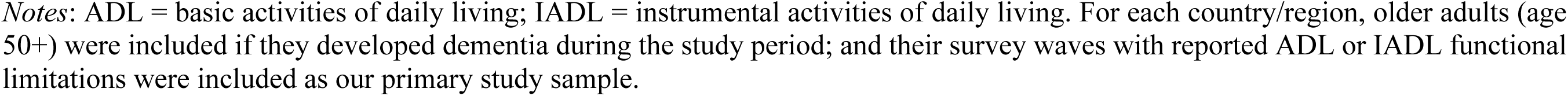
Flow chart of study design

### 2.2 Dementia Assessment

All older adults included in the sample reported functional limitations (as defined below) and developed dementia during the study period. Dementia status was determined using validated criteria for HRS-family studies.^27–30^. For HRS, we employed a well-established algorithm, classifying participants as PLWD if their 27-point cognition summary score was 6 or lower.^27,28^ The 27-point cognition scale comprises three cognitive tests: immediate and delayed word recall tests to measure memory (0-20 points), serial sevens subtraction test for working memory (0-5 points); and counting backwards test for speed of mental processing (0-2 points).

For ELSA, SHARE, and CHARLS, we used an alternative method since the above algorithm was specifically designed for HRS samples^27,28^ and may not be valid for other cohorts.^29,30^ Individuals were classified as PLWD if their cognition summary scores were 1.5 standard deviations (SDs) below the mean of the population stratified by education levels.^29,30^ As backward counting was not assessed, we used a 25-point cognition scale with the same word recall tests (0-20 points) and serial sevens test (0-5 points) as in HRS. Given the differences in cognitive scales and algorithms between HRS and the other three surveys, we performed sensitivity analyses where participants’ cognitive status was all evaluated using the same 25-score scale and defined based on the 1.5 SDs threshold.

For each country/region, dementia status was assigned in each wave, and participants were included if they had developed dementia during the study periods. Proxy assessment of cognition was not considered to ensure comparability across surveys.

### 2.3 Functional Limitations and Absence of Care

Functional limitations were assessed based on ADLs and IADLs. ADLs included six items: dressing, walking across a room, bathing, eating, getting in and out of bed, toileting; and IADLs included five items: preparing hot meals, shopping for groceries, making phone calls, taking medications, and managing money.^11,13^ Each ADL/IADL item was similarly assessed in the HRS-family surveys. The participants were asked if they have any difficulty with each ADL/IADL because of a health or memory problem or not. This resulted in 11 binary indicators of limitations, one for each ADL/IADL, reflecting different aspects or domains of functional limitations. In this study, we measured the extent of functional limitations by the number of ADL/IADL limitations (sum scores of ADL/IADLs, range 0–11), ADL limitations (sum scores of ADLs, range 0-6), and IADL limitations (sum scores of ADLs, range 0-5). Definitions and measurements across surveys are presented in Supplementary Table S2.

When relevant, participants were asked if they received any care for their functional limitations, with separate questions for the types of limitations (e.g., ADLs vs. IADLs) and the types of care received (formal care vs. informal care). To assess the absence of care for functional limitations, we constructed binary variables to indicate if participants received no care at all for their reported ADLs (0/1), IADLs (0/1), and ADLs/IADLs (0/1) limitations. In other words, the absence of care (for ADLs) meant that the participants did not receive any care for any of their reported (ADL) limitations. Additionally, for each type of functional limitation, we differentiated between the formal care and informal care. Therefore, the absence of care was defined respectively for ADLs (including 3 binary variables: no care, no informal care, no formal care), IADLs (3 binary variables), and for ADL/IADL altogether (3 binary variables). Survey questions and their similarities and differences across surveys are presented in Supplementary Table S3 and availability of data and variables are provided in Supplementary Table S4.

### 2.4 Statistical Analyses

Descriptive statistics of the sociodemographic characteristics, ADL/IADL limitations, and care received were first estimated using pooled person-wave data for each country/region. Categorical variables were reported as number (percentage), and continuous/count variables were reported as mean (standard deviation, SD). The missingness of the variables was minimal (mostly <1%) as shown in Supplementary Table S5.

Wave-specific estimates for absence of care were obtained using fitted models. To account for differences in sample compositions across waves, we calculated estimates of absence of care after adjustment for age, sex and the number of ADLs (0-6) and IADLs (0-5). The change in outcomes over time was estimated using linear models by adding interview year and aforementioned covariates as the predictors.^31^ The interview year coefficient captures the adjusted annual percent change (AAPC) for binary outcomes (i.e., receiving no care for ADL/IADL). Survey weights were applied in the analyses to account for sampling design and study attrition.

To examine potential disparities and differences in the absence of care, we further stratified our samples into two groups by educational attainment (less vs. more educated, stratified by median levels of education) and current-wave living arrangement (living alone vs. not living alone). Adjusted estimates of absence of care were obtained for each subgroup using pooled person-wave data, and Mann-Whitney-Wilcoxon rank-sum tests were used to test the distributional differences in adjusted values between subsamples.

STATA (version 17.0) was used to perform the analyses and all tests were two-sided with alpha level of 0.05 for statistical significance. The study followed the Strengthening the Reporting of Observational Studies in Epidemiology (STROBE) reporting guideline.

## 3. Results

### 3.1 Sample Characteristics

**Table 1** displays the sample characteristics of older adults with ADL/IADL limitations who developed dementia during the study periods. The average age (SD) of study samples ranged from 68.8 (9.1) years in CHARLS to 78.7 (9.4) years in SHARE; with notable cross-country/region differences in sociodemographic characteristics. Specifically, ELSA had the highest proportion of person-waves for living alone (42.4%), whereas CHARLS had the lowest proportion (10.2%). Additionally, 44.7% of person-waves in the HRS received at least secondary education, as compared to 5.5% in CHARLS.

**Table 1.**
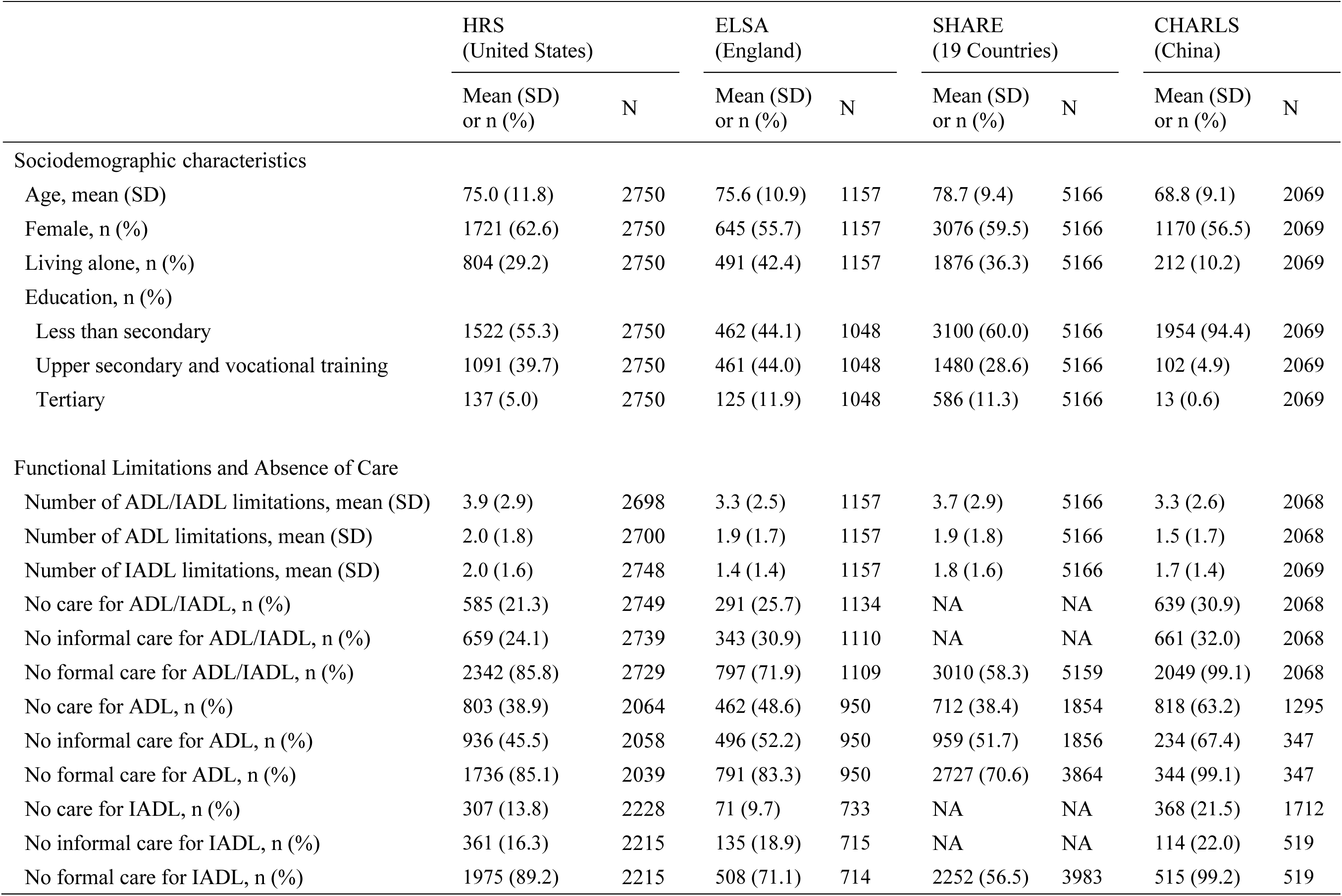
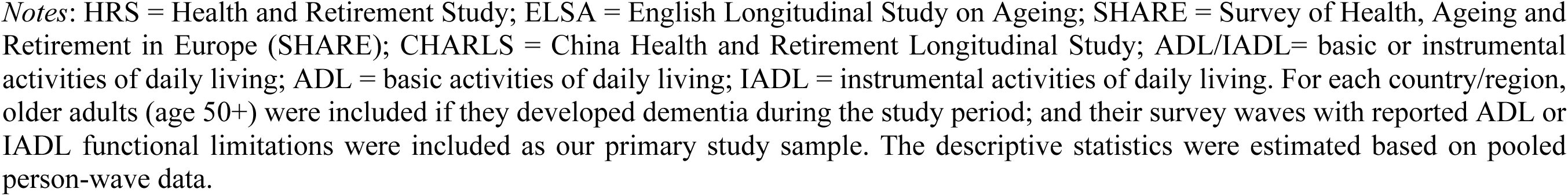
Characteristics of the study sample in the HRS, ELSA, SHARE, and CHARLS.

|  | HRS<br>(United States) |  | ELSA<br>(England) |  | SHARE<br>(19 Countries) |  | CHARLS<br>(China) |  |
| --- | --- | --- | --- | --- | --- | --- | --- | --- |
|  | Mean (SD)<br>or n (%) | N | Mean (SD)<br>or n (%) | N | Mean (SD)<br>or n (%) | N | Mean (SD)<br>or n (%) | N |
| Sociodemographic characteristics |  |  |  |  |  |  |  |  |
| Age, mean (SD) | 75.0 (11.8) | 2750 | 75.6 (10.9) | 1157 | 78.7 (9.4) | 5166 | 68.8 (9.1) | 2069 |
| Female, n (%) | 1721 (62.6) | 2750 | 645 (55.7) | 1157 | 3076 (59.5) | 5166 | 1170 (56.5) | 2069 |
| Living alone, n (%) | 804 (29.2) | 2750 | 491 (42.4) | 1157 | 1876 (36.3) | 5166 | 212 (10.2) | 2069 |
| Education, n (%) |  |  |  |  |  |  |  |  |
| Less than secondary | 1522 (55.3) | 2750 | 462 (44.1) | 1048 | 3100 (60.0) | 5166 | 1954 (94.4) | 2069 |
| Upper secondary and vocational training | 1091 (39.7) | 2750 | 461 (44.0) | 1048 | 1480 (28.6) | 5166 | 102 (4.9) | 2069 |
| Tertiary | 137 (5.0) | 2750 | 125 (11.9) | 1048 | 586 (11.3) | 5166 | 13 (0.6) | 2069 |
| Functional Limitations and Absence of Care |  |  |  |  |  |  |  |  |
| Number of ADL/IADL limitations, mean (SD) | 3.9 (2.9) | 2698 | 3.3 (2.5) | 1157 | 3.7 (2.9) | 5166 | 3.3 (2.6) | 2068 |
| Number of ADL limitations, mean (SD) | 2.0 (1.8) | 2700 | 1.9 (1.7) | 1157 | 1.9 (1.8) | 5166 | 1.5 (1.7) | 2068 |
| Number of IADL limitations, mean (SD) | 2.0 (1.6) | 2748 | 1.4 (1.4) | 1157 | 1.8 (1.6) | 5166 | 1.7 (1.4) | 2069 |
| No care for ADL/IADL, n (%) | 585 (21.3) | 2749 | 291 (25.7) | 1134 | NA | NA | 639 (30.9) | 2068 |
| No informal care for ADL/IADL, n (%) | 659 (24.1) | 2739 | 343 (30.9) | 1110 | NA | NA | 661 (32.0) | 2068 |
| No formal care for ADL/IADL, n (%) | 2342 (85.8) | 2729 | 797 (71.9) | 1109 | 3010 (58.3) | 5159 | 2049 (99.1) | 2068 |
| No care for ADL, n (%) | 803 (38.9) | 2064 | 462 (48.6) | 950 | 712 (38.4) | 1854 | 818 (63.2) | 1295 |
| No informal care for ADL, n (%) | 936 (45.5) | 2058 | 496 (52.2) | 950 | 959 (51.7) | 1856 | 234 (67.4) | 347 |
| No formal care for ADL, n (%) | 1736 (85.1) | 2039 | 791 (83.3) | 950 | 2727 (70.6) | 3864 | 344 (99.1) | 347 |
| No care for IADL, n (%) | 307 (13.8) | 2228 | 71 (9.7) | 733 | NA | NA | 368 (21.5) | 1712 |
| No informal care for IADL, n (%) | 361 (16.3) | 2215 | 135 (18.9) | 715 | NA | NA | 114 (22.0) | 519 |
| No formal care for IADL, n (%) | 1975 (89.2) | 2215 | 508 (71.1) | 714 | 2252 (56.5) | 3983 | 515 (99.2) | 519 |

*Notes:* HRS = Health and Retirement Study; ELSA = English Longitudinal Study on Ageing; SHARE = Survey of Health, Ageing and Retirement in Europe (SHARE); CHARLS = China Health and Retirement Longitudinal Study; ADL/IADL= basic or instrumental activities of daily living; ADL = basic activities of daily living; IADL = instrumental activities of daily living. For each country/region, older adults (age 50+) were included if they developed dementia during the study period; and their survey waves with reported ADL or IADL functional limitations were included as our primary study sample. The descriptive statistics were estimated based on pooled person-wave data.

The extent of ADL/IADL limitations and absence of care varied widely across countries/regions but showed some common patterns. The mean number of ADL/IADL limitations was consistently high across countries/regions, with approximately 3.3 in ELSA and CHARLS, 3.7 in SHARE, and 3.9 in the HRS. The prevalence of receiving no care for ADL/IADL limitations was at least 20% in all countries/regions, ranging from 21.3% in the US to 30.9% in China. Moreover, the absence of care was more pronounced for ADLs than IADLs. Notably, 48.6% and 63.2% of the samples in ELSA and CHARLS received no care at all for ADLs, and the proportions were also high in the HRS (38.9%) and SHARE (38.4%). Regarding the types of care, the prevalence of receiving no formal care for ADL/IADL limitations (ranging from 58.3% in SHARE to 99.1% in CHARLS) was much higher than the prevalence of receiving no informal care (ranging from 24.1% in the HRS to 32.0% in CHARLS). The patterns were similar for ADLs and IADLs.

### 3.2 Trends in the Absence of Care for Functional Limitations

**Figure 2** depicts the trends in the proportion of people with dementia who received no care for their ADL/IADL limitations by country/region, functional limitation type, and care type. Notably, almost all of these trends remained relatively stable (almost all P-values for AAPCs > 0.05). During the study period, at least 20% of PLWD had no care for their ADL/IADL limitations (Panel A). Specifically, about 34%-66% of individuals who developed dementia received no care for ADL limitations (Panel D), which is higher than the proportion receiving no care for IADL limitations (ranges from approximately 7% to 23%, Panel G) across all countries/regions from 2012 to 2018.

**Figure 2.**
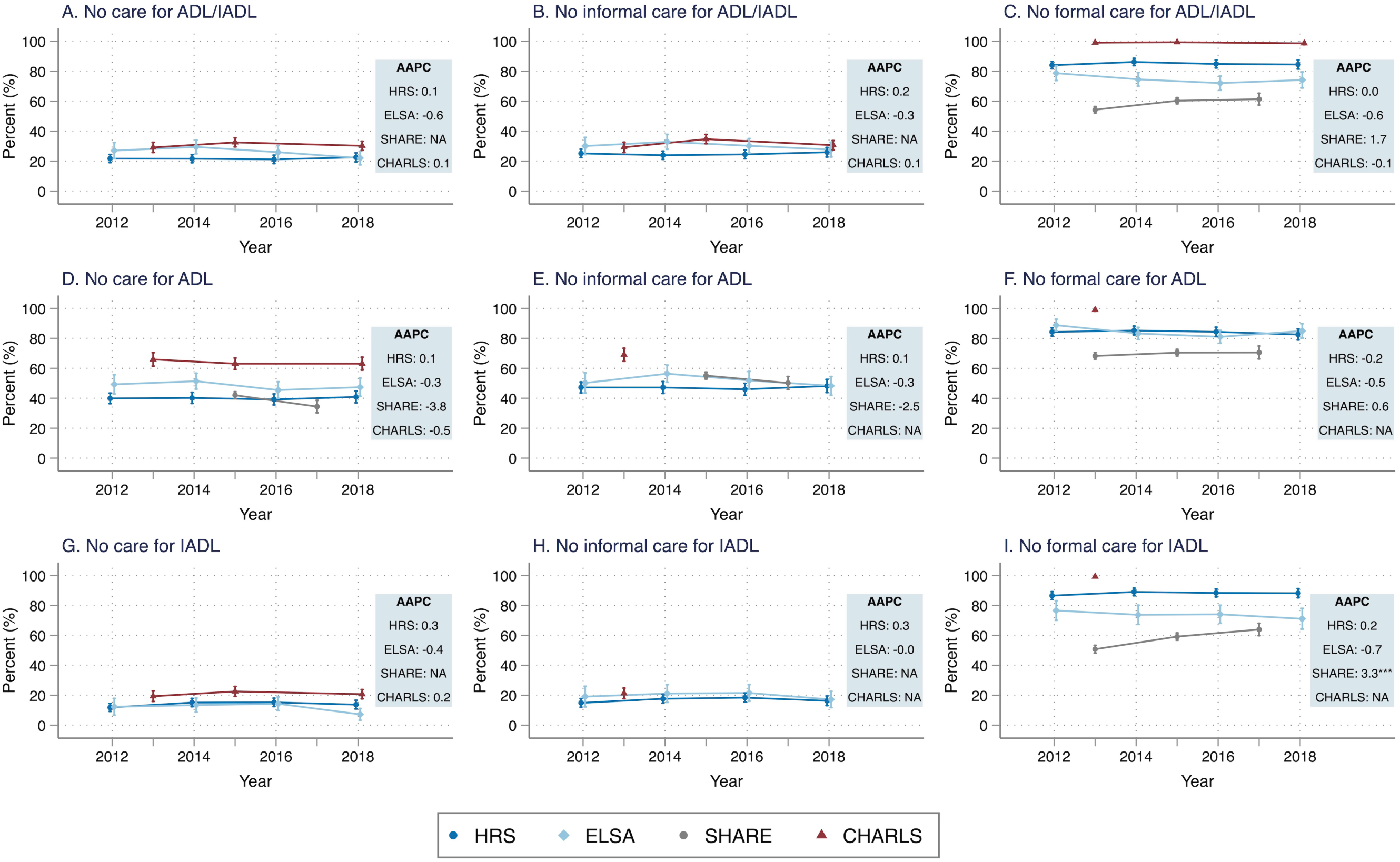
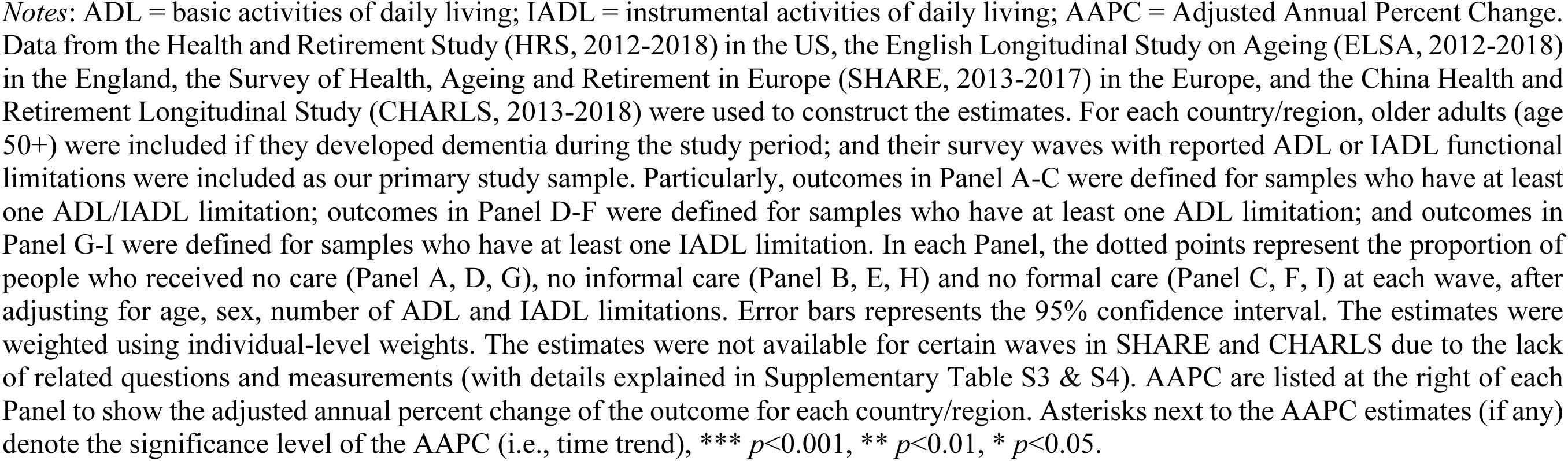
Trends in the proportion of persons receiving no care for ADL and IADL functional limitations among persons with dementia and functional limitations in the HRS, ELSA, SHARE, and CHARLS (2012-2018)

**Figure 2** also shows that the absence of care was more pronounced for formal care than informal care in all countries/regions throughout the study period, although both types of care exhibited high levels of absence (Panel B vs C, Panel E vs F, and Panel H vs I). Around 24%-35% of people with dementia received no informal care for their ADL/IADL limitations across countries/regions (Panel B), which is lower than the proportion receiving no formal care for their ADL/IADL limitations (ranging from 54% in SHARE to nearly 100% in CHARLS, Panel C). All four countries/regions showed minimal change in these proportions over the study period. Our sensitivity analyses supported the observed patterns, demonstrating that the estimates and trends were robust to the cognitive scale and algorithms used to classify dementia cases (Supplementary Figure S1).

### 3.3 Differences in the Absence of Care by Education and Living Arrangement

We further examined the differences in the absence of care for ADL/IADL limitations stratified by educational attainment and current-wave living arrangement. **Figure 3** reveals that less-educated individuals mostly reported a significantly higher proportion of not receiving formal care compared to more-educated individuals (Panel C, F, I). The differences in this proportion between the two groups were notable, with SHARE (13-16%) being the strongest and CHARLS being the lowest (1-2%) (Panel C, F, I). In contrast, there was no significant difference in the proportion of not receiving informal care for ADL/IADL limitations by educational attainment, except in ELSA where less-educated individuals reported lower proportion of receiving no informal care than their more-educated counterparts (Panel B, E, H). Overall, the absence of any care for ADL/IADL limitations was similar between more-educated and less-educated individuals in the HRS and SHARE, slightly more severe among more-educated individuals in ELSA, and among less-educated individuals in CHARLS (Panel A, D, G).

**Figure 3.**
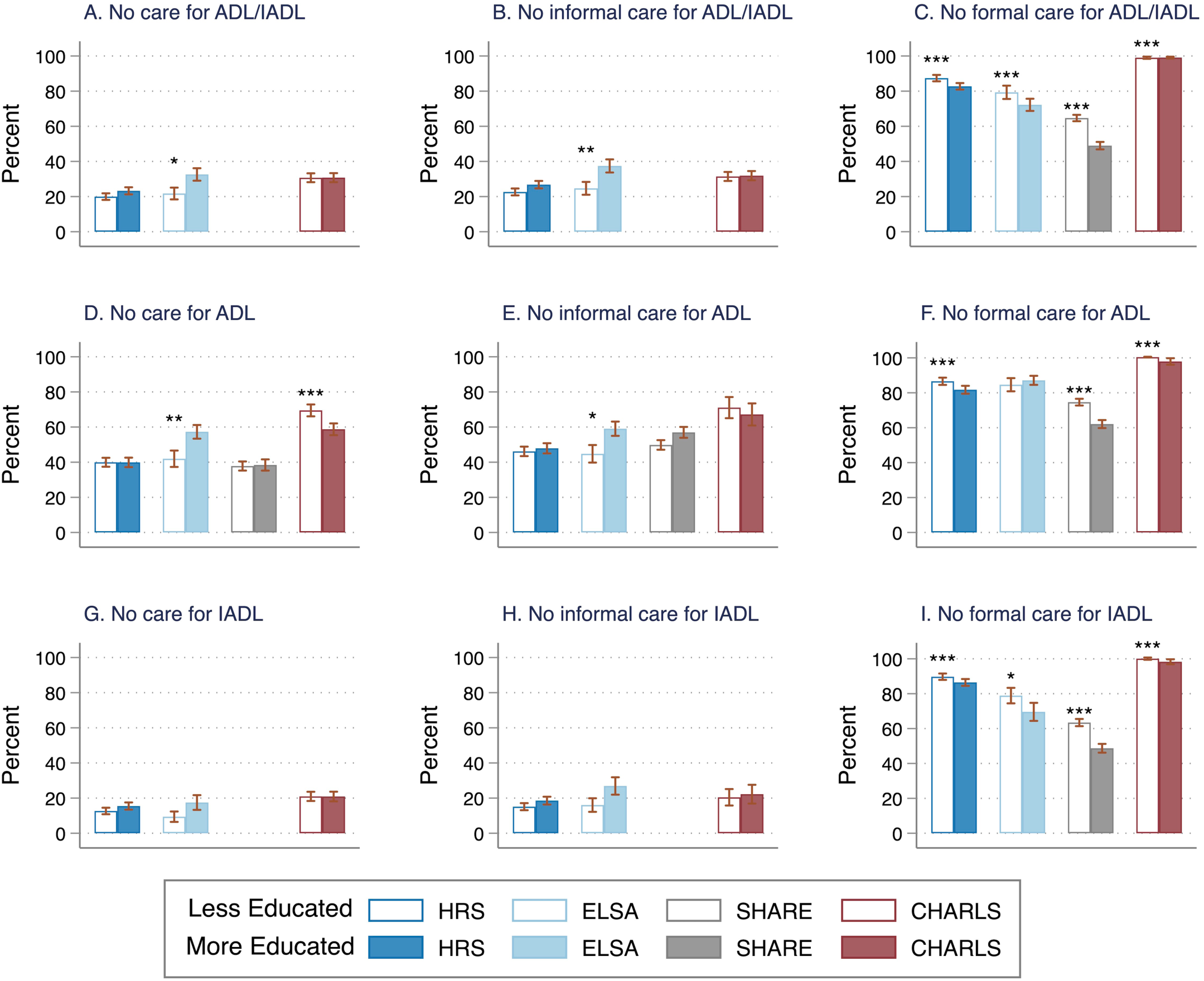
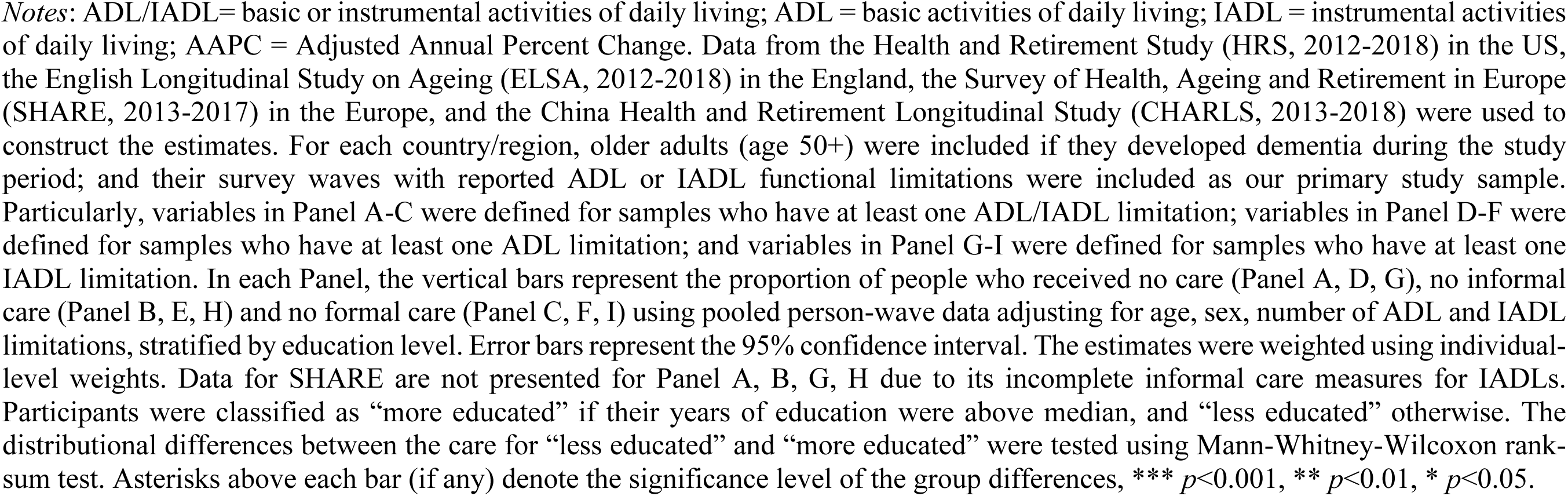
Proportion of persons receiving no care for ADL and IADL functional limitations among persons with dementia and functional limitations in the HRS, ELSA, SHARE, and CHARLS, stratified by educational attainment

Figure 4 showed that, for individuals with dementia living alone, the proportion receiving no informal care for ADL/IADL limitations was significantly higher than those living with others (Panel B, E, H). The difference ranged from 4% to 28% (Panel B, E, H). While the proportion of receiving no formal care for ADL/IADL limitations appeared to be lower among older adults living alone, especially in HRS, ELSA and SHARE (Panel C, F, I), they overall exhibited higher proportions receiving no care at all for ADL/IADL limitations compared to those living with others (Panel A, D, G). These patterns were similarly observed for both ADL and IADL limitations. Notably, the disparities in the absence of care between the two groups (living alone vs. not living alone) were substantial in HRS, ELSA and CHARLS, and the difference is as high as 21% (Panel A, D, G).

**Figure 4.**
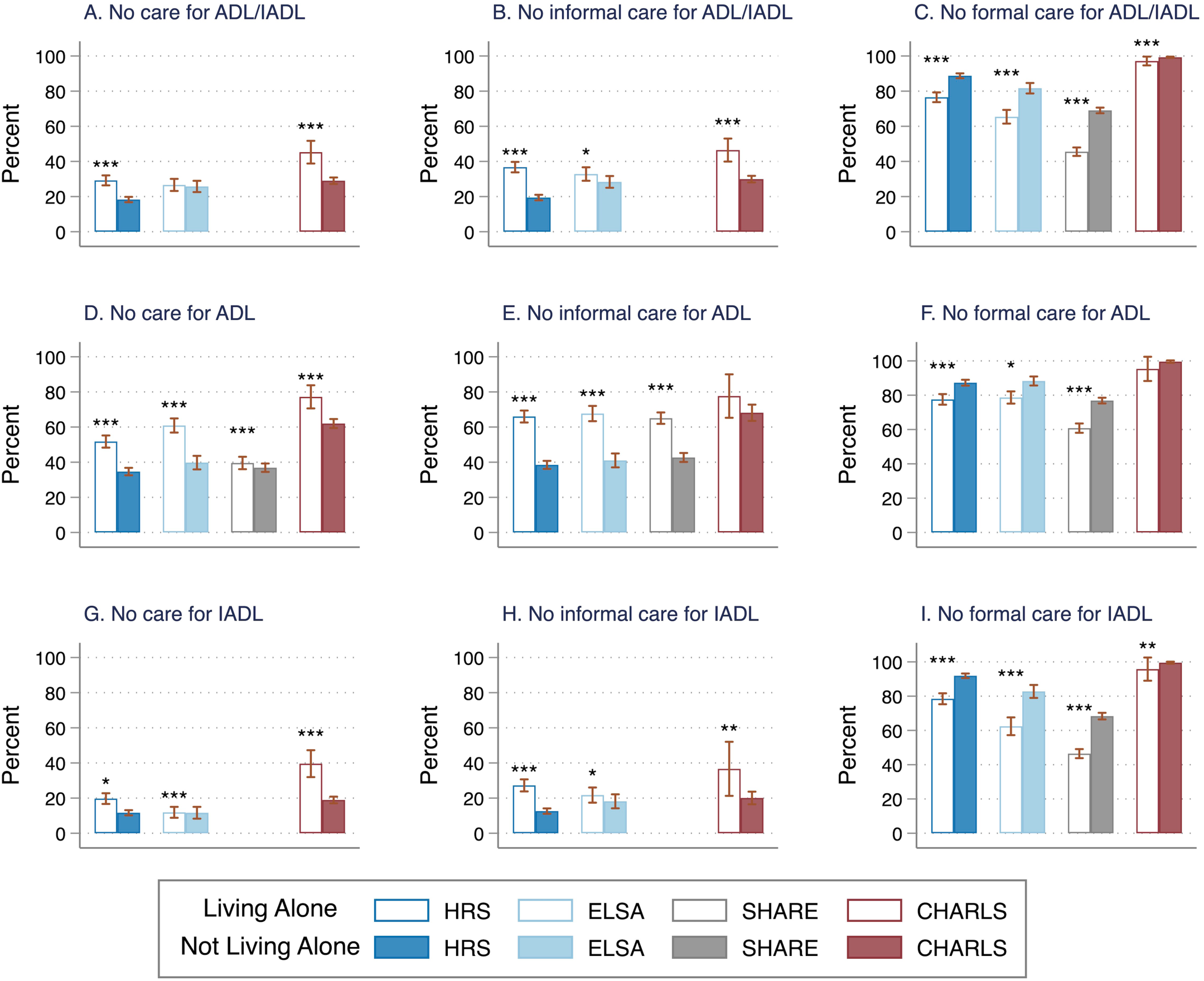
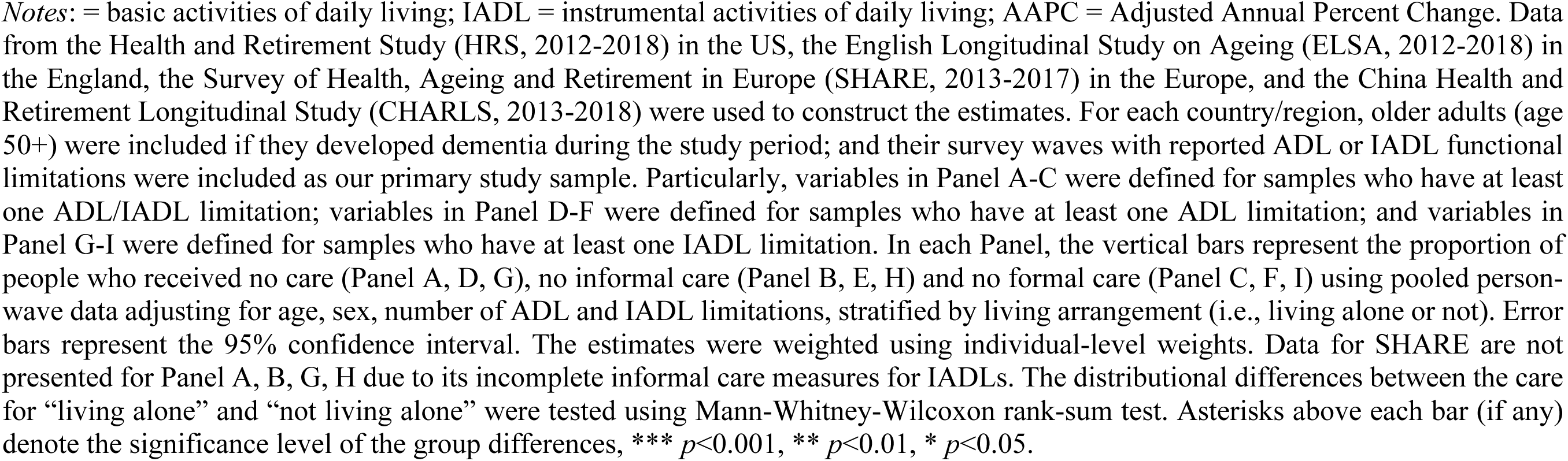
Proportion of persons receiving no care for ADL and IADL functional limitations among persons with dementia and functional limitations in the HRS, ELSA, SHARE, and CHARLS, stratified by living arrangement

## 4. Discussion

Using harmonized longitudinal surveys from the United States (HRS), England (ELSA), European countries and Israel (SHARE), and China (CHARLS), we presented the first comparative evidence on the global trends in the absence of care for PLWD with functional limitations. Our findings reveal two concerning facts: firstly, at least 1 in 5 PLWD across both developing and developed countries received no care for their ADL/IADL limitations; and secondly, this absence of care has not improved over time. Our analysis indicates that these trends hold true for both ADL and IADL limitations as well as for informal and formal care. Moreover, our results reveal that approximately 2 in 5 PLWD received no informal care for their ADL limitations and at least 3 in 5 (approaching 100% in China) received no formal care for their ADL limitations. Our findings highlight the significant absence of care for vulnerable populations, particularly among PLWD who were less educated or living alone.

Our analysis of cross-country/region data highlights a concerning and persistent gap in caregiving at a global scale for PLWD with functional limitations, a considerable proportion of whom received no care over time. The absence of care has been associated with adverse consequences such as anxiety, depression, increased risk of emergency room visits, hospitalization, nursing home admission, and premature death.^32–34^ PLWD with functional limitations are highly dependent on caregivers and are at even greater risk of experiencing the adverse consequences resulting from the absence of care.^35^ Whereas previous studies have pointed to the need for providers and policymakers to improve their efforts on ensuring adequate care provision for high-need patient populations, our evidence shows that the healthcare systems in both developing and developed countries in our sample failed to meet the care needs of their vulnerable constituents.

Although medical care for dementia is relatively well-insured in the US, long-term care (LTC) insurance coverage is limited and incomplete, leaving many with unmet care needs.^36^ Additionally, the US is facing a shortage of LTC workforce in nursing, home health, and personal care, posing challenges in providing adequate formal care services for PLWD. Furthermore, lack of nationwide paid caregiving leave and job protection for employees in the US makes it especially difficult for working caregivers to provide necessary care for PLWD with high caregiving demand. England and European countries have greater provision of public services and various forms of LTC facilities than other nations, but still lack services for people with special needs, such as PWLD.^37^ In China, the gap in LTC gap is much more pronounced than in developed countries. The formal LTC system is still in its early stages and faces significant challenges, including shortages of LTC facilities, workforce, and public financing.^38^ Despite the promotion of pilot LTC programs in China since 2016, the capacity of the LTC system remains insufficient. Importantly, few policies and measures have been developed for PLWD who require special attention,^39,40^ and most community-living PLWD depend on their family members for informal caregiving, with these caregivers receiving little support to alleviate care burden.

The persistent gap in caregiving for PLWD experiencing functional limitations across countries may be attributed to global population aging, the increasing burden of chronic diseases, and a decreased availability of children to provide care for aging parents with dementia.^13,41,42^ Furthermore, stigma related to functional limitations and dementia may discourage those affected from seeking care, which can further widen the gap in caregiving.^43^ As a result, caregiving for PLWD with functional limitations becomes increasingly complex and challenging over time, highlighting the need for continuous and targeted efforts to improve care provision for this population in both developing and developed countries.

Moreover, our study reveals a greater degree of absence of care for ADLs than IADLs among PLWD, which is consistent with previous research on older populations in the US, England and China^44–46^. Caregivers often face significant challenges in providing care with personal and mobility-related activities (i.e., ADLs), as noted in existing literature.^39,46^ Therefore, policy interventions and public programs should prioritize the provision of resources and support to caregivers to meet the basic care needs of PLWD experiencing functional limitations, especially those with ADL limitations.

Our findings also indicate a higher absence of formal care than informal care among PLWD who experience functional limitations across countries and regions. This is not surprising given that informal care is the primary source of care for PLWD and can serve as a substitute for formal care.^47,48^ However, the significant proportion of PLWD who receive no formal care for ADL/IADL limitations across countries/regions is concerning given their tremendous needs for professional services.^5,39,43^ As previously discussed, the absence of formal care among PLWD may reflect an inadequate provision of formal care services and obstacles hindering PLWD’s access to such services. The appropriate use of additional paid services or formal care could improve physical functioning for care recipients.^49^ Assistance from paid caregivers or institutions could also help manage complex medical or support needs of patients and alleviate the caregiving burden for family members. Therefore, achieving a balance between the use of formal and informal care services is crucial in improving health outcomes for PLWD and their caregivers.

Lastly, our study highlights the notable disparities in different types of absence of care for PLWD with functional limitations, which can be especially salient among vulnerable populations facing difficulties in accessing care and resources.^42,50^ Our findings demonstrate that PLWD with lower levels of education had greater absence of formal care, and those who lived alone had greater absence of informal care and overall care compared to their counterparts. These individuals likely have the greatest functional limitations,^51,52^ thus the highest care needs. Therefore, it is imperative for policymakers to prioritize ensuring adequate care provision for the most vulnerable groups. This involves enhancing their access to LTC, expanding the range of LTC options available to them, and empowering PLWD to make well-informed decisions about their care choices.

### Limitations

Although our study stands out for harmonizing the data to provide comparative evidence on a global scale for the absence of care among PLWD with functional limitations, it has several limitations. First, we relied on both self-reported and proxy-reported measures of ADL/IADL limitations, which can be subject to reporting errors in terms of the extent of difficulties and support received. Second, we used cognitive scales to identify PLWD instead of clinicians, which may lead to classification errors, especially for those around the threshold of dementia. However, our sensitivity analysis confirmed the consistency of our results. Finally, this study focused on assessing the prevalence of absence of care for PLWD who have functional limitations, as well as the trajectories of such absence over time across countries/regions. The study did not estimate the quality of care received or the extent to which the individuals were satisfied with their care. Future studies should collect more data to fill the gap in existing data sources.

### Conclusion

Our study provides the first comparable evidence that a large proportion of PLWD with functional limitations receive no formal or informal care, and the shortage of care did not improve over time in both developing and developed countries. Formal care is more lacking than informal care, and ADLs are more affected than IADLs. Furthermore, we found notable disparities in the lack of formal care by educational attainment and in the lack of informal and overall care by living arrangement. These findings apply to all studied countries and regions. Our study highlights the urgent need for policy and practice improvements to enhance care provision for ADL/IADL limitations among PLWD and to allocate targeted LTC resources for those with socio-economic disadvantages worldwide.

## Data Availability

All data produced in the present study are available upon reasonable request to the authors.

## Acknowledgements

XC acknowledges research funding from the U.S. National Institute on Aging (R01AG077529; K01AG053408). TMG is supported by P30AG021342 from the U.S. National Institute on Aging. XC and YQ report serving under a contract with the World Bank on leading a report that assesses demand for and supply of home-based support for older adults with disabilities in 31 countries. SC’s research was supported by the PENDA, funded by the UK Foreign, Commonwealth and Development Office. The views expressed are those of the authors and not necessarily those of the funders. The funders had no role in the design and conduct of the study; collection, management, analysis, and interpretation of the data; preparation, review, or approval of the manuscript; or decision to submit the manuscript for publication. The authors acknowledge research assistance of Zexuan Yu at Brown University School of Public Health.

## Figures and Tables

**Figure S1.**
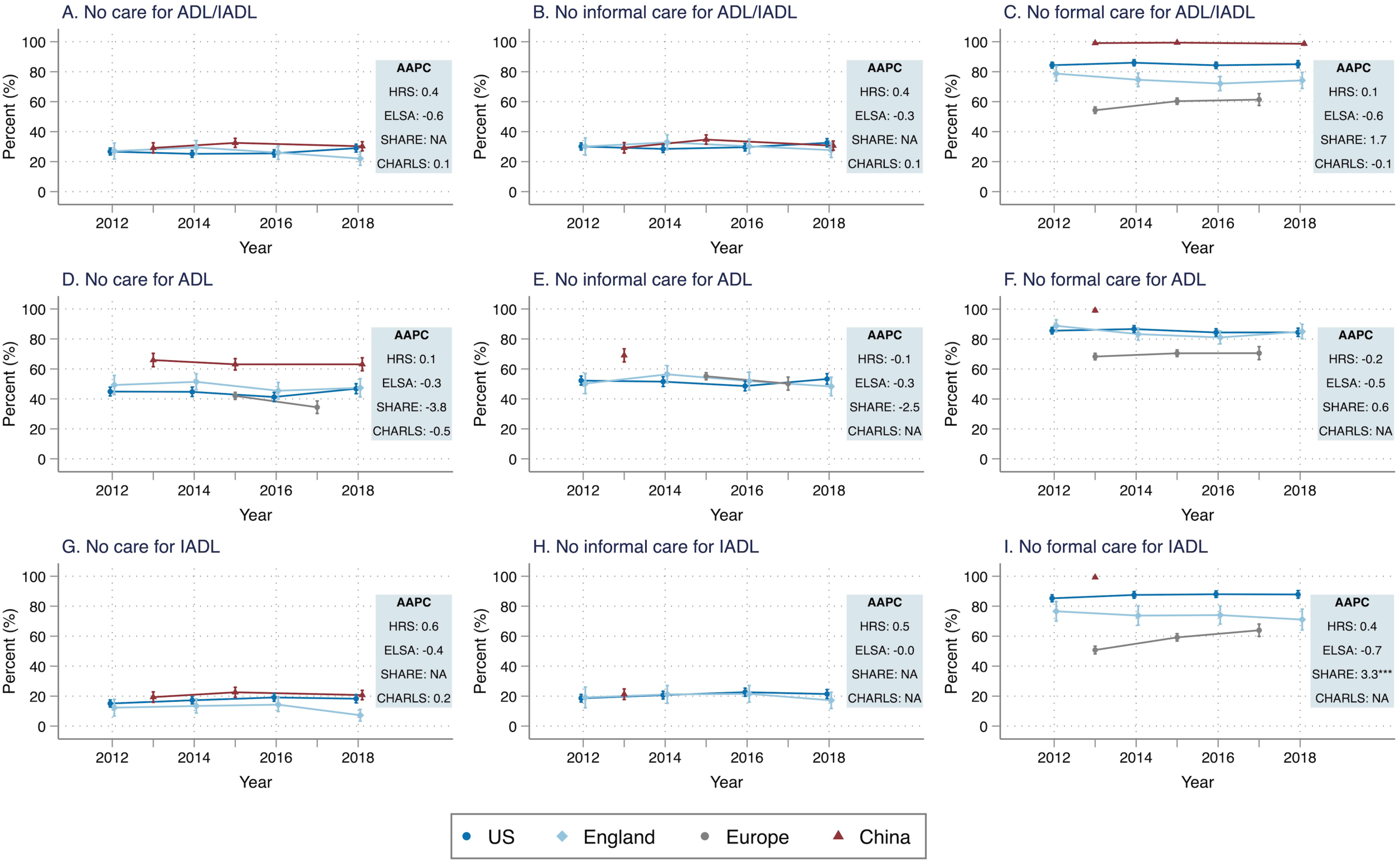
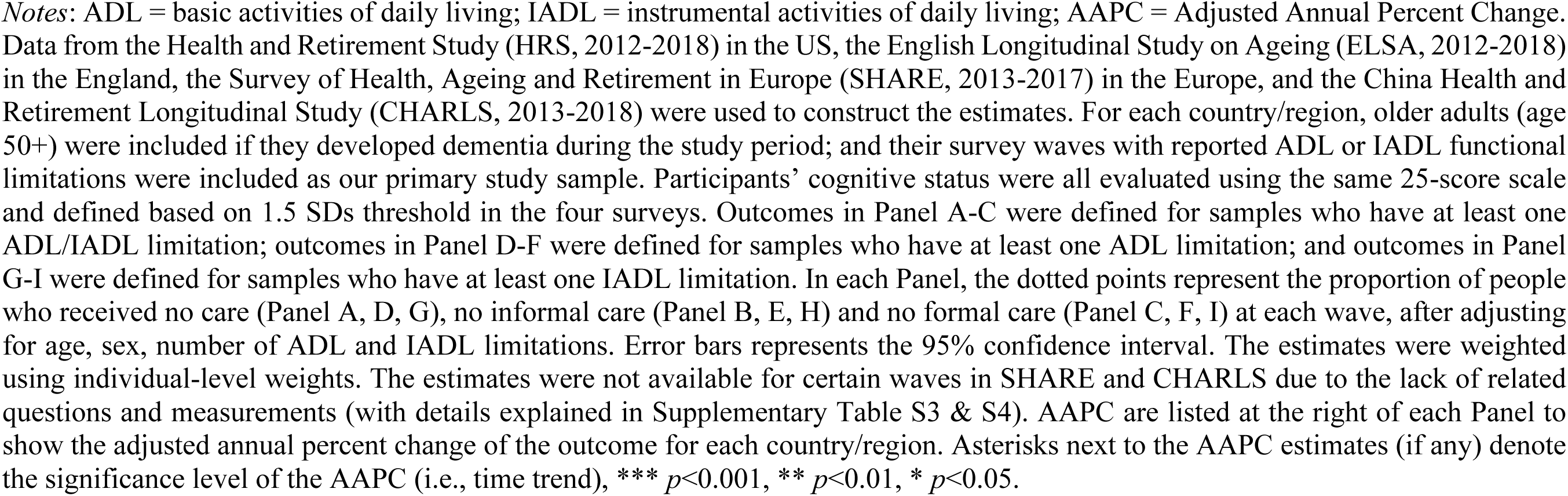
Trends in the proportion of people receiving no care for ADL and IADL functional limitations among persons with dementia and functional limitations in the HRS, ELSA, SHARE, and CHARLS, 2012-2018 (sensitivity analysis: participants’ cognitive status were all evaluated using the same 25-score scale and defined based on 1.5 SDs threshold in the four surveys)

**Supplementary Table S1.** Countries included in each study.

| Study | Countries |
| --- | --- |
| HRS | United States |
| ELSA | England |
| SHARE | Austria, Germany, Sweden, Netherland,<br>Spain, Italy, France, Denmark, Greece,<br>Switzerland, Belgium, Israel, Czech<br>Republic, Poland, Luxembourg, Portugal,<br>Slovenia, Estonia, Croatia |
| CHARLS | China |

*Notes:* HRS = Health and Retirement Study; ELSA = English Longitudinal Study on Ageing; SHARE = Survey of Health, Ageing and Retirement in Europe (SHARE); CHARLS = China Health and Retirement Longitudinal Study.

**Supplementary Table S2.** Survey questions of ADL and IADL functional limitations in HRS, ELSA, SHARE, and CHARLS.

|  | HRS (2012-2018) | ELSA (2012-2018) | SHARE (2013-2017) | CHARLS (2013-2018) |
| --- | --- | --- | --- | --- |
| ADL limitation | Here are a few more everyday activities. Please tell me if you have any difficulty with these because of a physical, mental, emotional or memory problem. Again, exclude any difficulties you expect to last less than three months. Because of a health or memory problem do you have any difficulty with:<br>• Dressing, including putting on shoes and socks.<br>• Walking across a room.<br>• Bathing or showering.<br>• Eating, such as cutting up your food.<br>• Getting in or out of bed.<br>• Using the toilet, including getting up or down. | Here are a few more everyday activities. Please tell me if [<sup>^</sup>you have / [<sup>^</sup>name] has] any difficulty with these because of a physical, mental, emotional or memory problem. Again, exclude any difficulties you expect to last less than three months. Because of a health or memory problem, [<sup>^</sup>do you /does he /does she] have difficulty doing any of the activities on this card?<br>• Dressing, including putting on shoes and socks.<br>• Walking across a room.<br>• Bathing or showering.<br>• Eating, such as cutting up [<sup>^</sup>your / his / her] food.<br>• Getting in or out of bed.<br>• Using the toilet, including getting up or down. | Here are a few more everyday activities. Please tell me if you have any difficulty with these because of a physical, mental, emotional or memory problem. Again, exclude any difficulties you expect to last less than three months.<br>• Dressing, including putting on shoes and socks.<br>• Walking across a room.<br>• Bathing or showering.<br>• Eating, such as cutting up your food.<br>• Getting in or out of bed.<br>• Using the toilet, including getting up or down. | Here are a few more everyday activities. Please tell me if you have any difficulty with these because of a physical, mental, emotional or memory problem. Again, exclude any difficulties you expect to last less than three months. Because of a health or memory problem do you have any difficulty with:<br>• Dressing? Dressing includes taking clothes out from a closet, putting them on, buttoning up, and fastening a belt.<br>• Bathing or showering<br>• Eating, such as cutting up your food? (Definition: By eating, we mean eating food by oneself when it is ready)<br>• Getting in or out of bed.<br>• Using the toilet, including getting up or down.<br>• Controlling urination and defecation? |
| IADL limitation | Here are a few other activities which some people have difficulty with because of a physical, mental, emotional, or memory problem. Please tell me whether you have any | Asked together with ADL (See the leading question above)<br>• Preparing a hot meal.<br>• Shopping for groceries. | Asked together with ADL (See the leading question above)<br>• Preparing a hot meal.<br>• Shopping for groceries. | Asked together with ADL (See the leading question above)<br>• Preparing hot meals? (Definition: By preparing hot meals, we mean preparing |
|  | difficulty with each activity I name. If you don't do the activity at all, just tell me so. Exclude any difficulties that you expect to last less than three months.<br>• Preparing a hot meal.<br>• Shopping for groceries.<br>• Making phone calls.<br>• Taking medications.<br>• Managing your money, such as paying bills and keeping track of expenses. | • Making telephone calls.<br>• Taking medications.<br>• Managing money, such as paying bills and keeping track of expenses. | • Making telephone calls.<br>• Taking medications.<br>• Managing money, such as paying bills and keeping track of expenses. | ingredients, cooking, and serving food)<br>• Shopping for groceries? By shopping, we mean deciding what to buy and paying for it.<br>• Making phone calls?<br>• Taking medications? By taking medications, we mean taking the right portion of medication right on time.<br>• Managing your money, such as paying your bills, keeping track of expenses, or managing assets? |

*Notes:* The number of ADL/IADL limitations were constructed as the sum of the ADL and IADL limitations.

**Supplementary Table S3.** Survey questions of care received for ADL and IADL functional limitations in HRS, ELSA, SHARE, and CHARLS.

|  | HRS (2012-2018) | ELSA (2012-2018) | SHARE (2013-2017) | CHARLS (2013-2018) |
| --- | --- | --- | --- | --- |
| Received any care for ADL limitations | If the respondent reports having difficulty with any ADL items, then they are asked immediately whether someone helps them with such activity (hence the definitions of each items have been clearly explained in the previous question). The questions are listed below:<br>• Does anyone ever help you dress?<br>• Does anyone ever help you get across a room<br>• Does anyone ever help you bathe?<br>• Does anyone ever help you eat<br>• Does anyone ever help you get in or out of bed?<br>• Does anyone ever help you use the toilet? | Starting in Wave 6 (2012-2018), if the respondent reports having difficulty with at least one ADL activity, then they are asked whether someone helps them with each activity. The questions are listed below:<br>• [^Have/Has] [^you/[^name]] received help from anyone with dressing, including putting on shoes and socks, in the last month?<br>• [^Have/Has] [^you/[^name]] received help from anyone with walking across a room, in the last month?<br>• [^Have/Has] [^you/[^name]] received help from anyone with bathing or showering, in the last month?<br>• [^Have/Has] [^you/[^name]] received help from anyone with eating, such as cutting up food, in the last month?<br>• [^Have/Has] [^you/[^name]] | Unlike HRS, ELSA, CHARLS, SHARE does not ask the respondents whether they receive help for each individual ADL items. Instead, SHARE asks if they receive informal helps from inside and outside the household respectively, and if they receive formal care at home.<br>For informal care received inside household, the types of care are assumed to be ADL-related helps.<br>• Is there someone living in this household who has helped you regularly during the last twelve months with personal care, such as washing, getting out of bed, or dressing?<br>For informal care received outside household, the types of care (ADL vs IADL) can only be identified since Wave 6 (2015-2017).<br>• Thinking about the last twelve months, has any family member from outside the household, any friend | If the respondent reports having difficulty with any ADL items, then they are asked immediately whether someone helps them with such activity (hence the definitions of each items have been clearly explained in the previous question). The questions are listed below:<br>• Does anyone ever help you dress?<br>• Does anyone ever help you bathe?<br>• Does anyone ever help you eat<br>• Does anyone ever help you get in or out of bed?<br>• Does anyone ever help you use the toilet? |
|  |  | received help from anyone with getting in or out of bed, in the last month?<br>• [^Have/Has] [^you/[^name]] received help from anyone with using the toilet, including getting up or down, in the last month? | or neighbour given you any kind of help listed on this card?<br>• Which types of help has this person provided in the last twelve months? [personal care, e.g., dressing, bathing or showering, eating, getting in or out of bed, using the toilet]<br>For formal care, the types of care (ADL vs IADL) are identifiable in all waves from 2013 to 2017.<br>• During the last twelve months, did you receive in your own home any professional or paid services listed on this card due to a physical, mental, emotional or memory problem? [Help with personal care, e.g., getting in and out of bed, dressing, bathing and showering]<br>Therefore, complete care for ADLs, including care received inside or outside the household and formal home care can only be measured in 2015- 2017. |  |
| Received any care for IADL limitations | If the respondent reports having difficulty with any IADL items, then they are asked immediately whether someone helps them with such activity (hence the definitions of each items have been clearly explained in the previous question). The questions are listed below:<br>• Does anyone help you prepare hot meals?<br>• Does anyone help you shop for groceries?<br>• Does anyone help you make telephone calls?<br>• Does anyone help you take medications?<br>• Does anyone help you manage your money? | Starting in Wave 6 (2012-2018), if the respondent reports having difficulty with at least one IADL activity, then they are asked whether someone helps them with each activity. The questions are listed below:<br>• [^Have/Has] [^you/[^name]] received help from anyone with shopping for groceries, in the last month?<br>• [^Have/Has] [^you/[^name]] received help from anyone with taking medications, in the last month?<br>• [^Have/Has] [^you/[^name]] received help from anyone with doing work around the house or garden, in the last month?<br>• [^Have/Has] [^you/[^name]] received help from anyone with managing money, such as paying bills and keeping track of expenses, in the last month? | Unlike HRS, ELSA, CHARLS, SHARE does not ask the respondents whether they receive help for each individual IADL items. Instead, SHARE asked if they receive informal helps from inside and outside the household respectively, and if they receive formal care at home.<br>For informal care received inside household, NO care is asked for IADL items.<br>For informal care received outside household, the types of care (ADL vs IADL) can only be identified since Wave 6 (2015-2017).<br>For formal care, the types of care (ADL vs IADL) are identifiable in all waves from 2013 to 2017.<br>Therefore, complete care for IADLs (including both informal and formal) can NOT be meaningfully measured in SHARE because informal care received from inside the household are not asked for IADLs. | If the respondent reports having difficulty with any IADL items, then they are asked immediately whether someone helps them with such activity (hence the definitions of each items have been clearly explained in the previous question). The questions are listed below:<br>• Does anyone help you do household chores?<br>• Does anyone help you prepare hot meals?<br>• Does anyone help you shop for groceries?<br>• Does anyone help you make telephone calls?<br>• Does anyone help you take medications?<br>• Does anyone help you manage your money? |
| Received any informal care for ADL limitations | If someone helps with any ADL activity, the respondents are asked for the relationships | Starting in Wave 6 (2012-2018), respondents who receive help are asked to separately list the | SHARE asks the respondents if they receive informal helps | In wave 2 (2013), the respondents who received help are asked for the relationship of |
|  | of up to 7 people who most often help them with ADLs (ADL items altogether). The questions are listed below:<br>Who most often helps you with [getting across a room/dressing /bathing/eating/getting (in/out of) bed/using the toilet]?<br>• Spouse or partner<br>• Son<br>• Stepson<br>• Spouse or partner of son<br>• Daughter<br>• Stepdaughter<br>• Spouse or partner of daughter<br>• Grandchild<br>• Father<br>• Father-in-law<br>• Mother<br>• Mother-in-law<br>• Brother<br>• Brother-in-law<br>• Sister<br>• Sister-in-law<br>• Other relative<br>• Other individual<br>• Former child-in-law<br>• Grandchild's spouse or partner | relationships for all the people who help according to the following groupings of ADL items: mobility (walking 100 yards, climbing several flights of stairs, climbing one flight of stairs, walking across a room, getting in or out of bed, using the toilet), bathing/showering or getting dressed, eating. Informal helpers include:<br>• Husband/Wife/Partner<br>• Son<br>• Daughter<br>• Grandchild<br>• Sister<br>• Brother<br>• Other relative<br>• Friend<br>• Neighbor | from inside and outside the household respectively. For informal care received inside household, the types of care are assumed to be ADL-related helps.<br>• Is there someone living in this household who has helped you regularly during the last twelve months with personal care, such as washing, getting out of bed, or dressing?<br>For informal care received outside household, the types of care (ADL vs IADL) can only be identified since Wave 6 (2015-2017).<br>• Thinking about the last twelve months, has any family member from outside the household, any friend or neighbour given you any kind of help listed on this card?<br>• Which types of help has this person provided in the last twelve months?<br>[personal care, e.g., dressing, bathing or showering, eating, getting in or out of bed, using the toilet] | up to 7 most often helpers for all ADL items altogether (but not for each). However, in wave 3-4 (2015-2018), the respondents who received help are asked for the most often helpers for ADL and IADL items altogether (but not separately for ADL items and IADL items). Therefore, helpers' types can only be identified for ADL items in 2013, but not in later waves. The questions in 2013 are listed below:<br>Who most often helps you with [dressing/bathing/eating/getting in out of bed/using the toilet]?<br>• Spouse<br>• Ex-spouse<br>• Mother<br>• Father<br>• Mother-in-law<br>• Farther-in-law<br>• Children [preload name]<br>• Sibling<br>• Sibling of spouse<br>• Brother-in-law, sister-in-law<br>• Grandson<br>• Granddaughter<br>• Other relative |
| Received any formal care for ADL limitations | Asked together with ADL informal helpers. See the detailed description above.<br>Who most often helps you with [getting across a room/dressing /bathing/eating/getting (in/out of) bed/using the toilet]?<br>• Nursing home<br>• Organization<br>• Employee of facility<br>• Paid helper | Asked together with ADL informal helpers. See the detailed description above. A list of formal caregivers include:<br>• Home care worker/ home help/ personal assistant;<br>• A member of the reablement / intermediate care staff team;<br>• Voluntary helper;<br>• Warden / Sheltered housing Manager;<br>• Cleaner;<br>• Council's handyman;<br>• Member of staff at the care/nursing home<br>• Other formal helper | For formal care, the types of care for ADLs are identifiable in all waves during 2013-2017.<br>• During the last twelve months, did you receive in your own home any professional or paid services listed on this card due to a physical, mental, emotional or memory problem? [Help with personal care, e.g., getting in and out of bed, dressing, bathing and showering] | Asked together with ADL informal helpers in 2013. See the detailed description above.<br>Who most often helps you with [dressing/bathing/eating/getting in out of bed/using the toilet]?<br>• Paid helper (such as nanny)<br>• Volunteer or employee of facility<br>• Nursing home |
| Received any informal care for IADL limitations | If someone helps with any ADL activity, the respondents are asked for the relationships of up to 7 people who most often help them with IADLs (IADL items altogether). The questions are listed below:<br>Who most often helps you with [prepare hot meals, /shop for groceries, /make telephone calls, /take medications]?<br>Who most often helps you manage your money?<br>• Spouse or partner<br>• Son<br>• Stepson<br>• Spouse or partner of son | Starting in Wave 6 (2012-2018), respondents who receive help are asked to separately list the relationships for all the people who help according to the following groupings of IADL items: shopping for groceries or doing work around the house or garden, taking medication, or managing money. Informal helpers include:<br>• Husband/Wife/Partner<br>• Son<br>• Daughter<br>• Grandchild<br>• Sister<br>• Brother<br>• Other relative<br>• Friend | SHARE asked if they receive informal helps from inside and outside the household respectively.<br>For informal care received inside household, NO care is asked for IADL items.<br>For informal care received outside household, the types of care (ADL vs IADL) can only be identified since Wave 6 (2015-2017).<br>• Thinking about the last twelve months, has any family member from outside the household, any friend or neighbour | In wave 2 (2013), the respondents who received help are asked for the relationship of up to 6 most often helpers for all IADL items altogether (but not for each). However, in wave 3-4 (2015-2018), the respondents who received help are asked for the most often helpers for ADL and IADL items altogether (but not separately for ADL items and IADL items). Therefore, helpers' types can only be identified for IADL items in 2013, but not in later waves. The questions in 2013 are listed below:<br>Who most often helps you with [doing household chores/preparing hot meals/shopping/making |
|  | • Daughter<br>• Stepdaughter<br>• Spouse or partner of daughter<br>• Grandchild<br>• Father<br>• Father-in-law<br>• Mother<br>• Mother-in-law<br>• Brother<br>• Brother-in-law<br>• Sister<br>• Sister-in-law<br>• Other relative<br>• Other individual<br>• Former child-in-law<br>• Grandchild's spouse or partner | • Neighbor | given you any kind of help listed on this card?<br>• Which types of help has this person provided in the last twelve months? [practical household help, e.g. with home repairs, gardening, transportation, shopping, household chores; Help with paperwork, such as filling out forms, settling financial or legal matters]<br>Therefore, informal care for IADLs can NOT be meaningfully measured in SHARE because informal care received from inside the household are not asked for IADLs. | telephone calls/taking medications]?<br>• Spouse<br>• Ex-spouse<br>• Mother<br>• Father<br>• Mother-in-law<br>• Farther-in-law<br>• Children [preload name]<br>• Sibling<br>• Sibling of spouse<br>• Brother-in-law, sister-in-law<br>• Grandson<br>• Granddaughter<br>• Other relative |
| Received any formal care for IADL limitations | Asked together with IADL informal helpers. See the detailed description above.<br>Who most often helps you with [prepare hot meals, /shop for groceries, /make telephone calls, /take medications]?<br>Who most often helps you manage your money?<br>• Nursing home<br>• Organization<br>• Employee of facility | Asked together with IADL informal helpers. See the detailed description above. A list of formal helpers include:<br>• Home care worker/ home help/ personal assistant;<br>• A member of the reablement / intermediate care staff team;<br>• Voluntary helper;<br>• Warden / Sheltered housing Manager; | For formal care, the types of care for IADLs are identifiable in all waves during 2013-2017.<br>• During the last twelve months, did you receive in your own home any professional or paid services listed on this card due to a physical, mental, emotional or memory problem? [Help with | Asked together with IADL informal helpers in 2013. See the detailed description above.<br>Who most often helps you with [doing household chores/preparing hot meals/shopping/making telephone calls/taking medications]<br>• Paid helper (such as nanny)<br>• Volunteer or employee of facility |
|  | • Paid helper | • Cleaner;<br>• Council's handyman;<br>• Member of staff at the care/nursing home<br>• Other formal helper | domestic tasks, e.g., cleaning, ironing, cooking; Help with meals, i.e., ready-made meals provided by a municipality or a private provider; Help with other activities, e.g., filling a drug dispenser] | • Nursing home |

*Notes:* The (informal/formal) care received for ADL/IADL altogether were mostly constructed based on the (informal/formal) care received for ADLs and IADLs separately. In some cases, such as CHARLS 2015-2018, the informal/formal care received for ADL/IADL were asked in one question for all ADL/IADL items but not for ADLs and IADLs separately; and the informal/formal care received were constructed based on that question.

**Supplementary Table S4.**
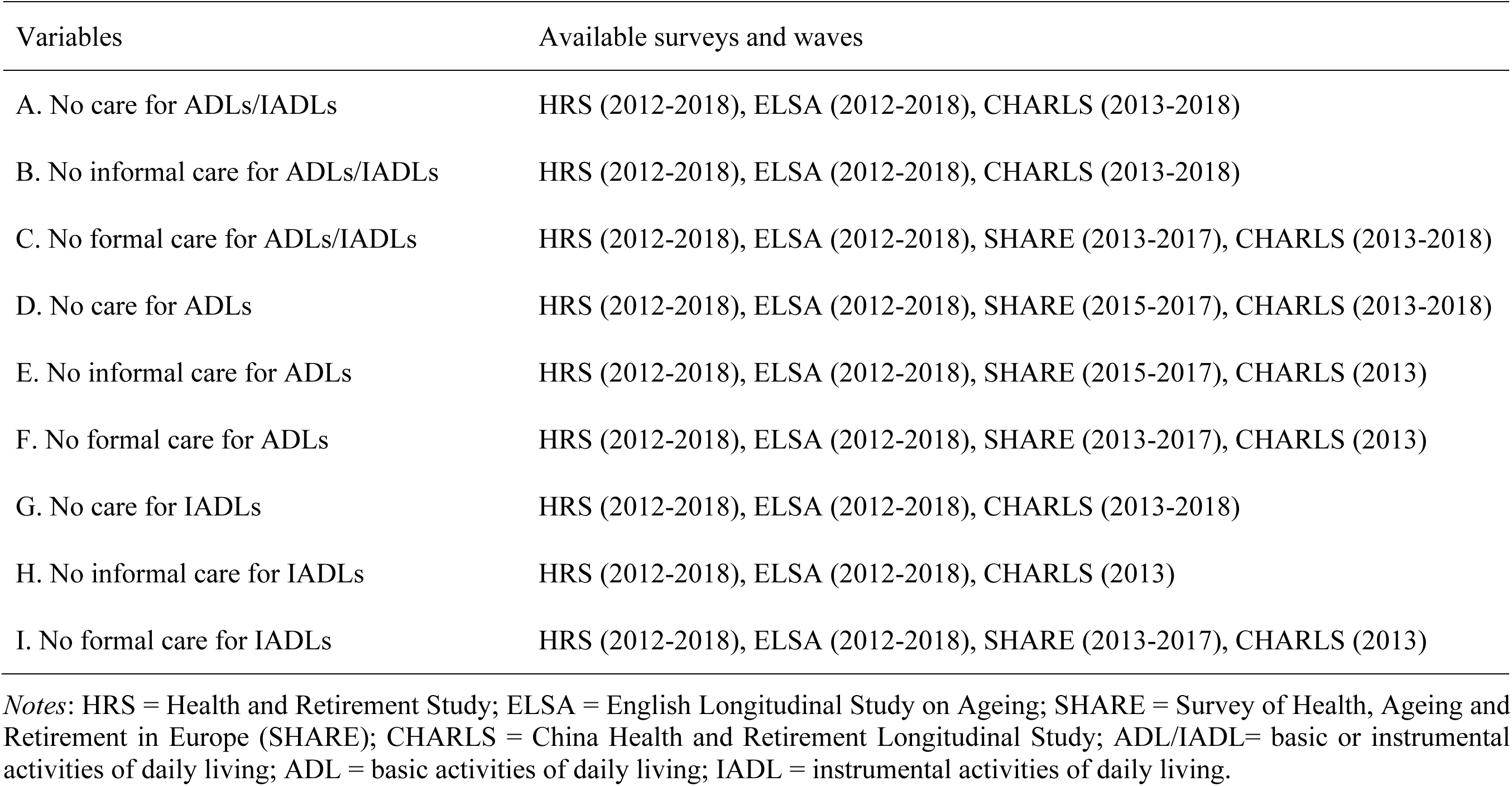
Availability of data (surveys and waves) for binary variables denoting absence of care.

**Supplementary Table S5.** Missingness of variables (i.e., n (%)) among all eligible samples who were asked the survey questions.

| Variables | HRS<br>(United States) |  | ELSA<br>(England) |  | SHARE<br>(19 Countries) |  | CHARLS<br>(China) |  |
| --- | --- | --- | --- | --- | --- | --- | --- | --- |
|  | n (%) | Total | n (%) | Total | n (%) | Total | n (%) | Total |
| Functional Limitations and Absence of Care |  |  |  |  |  |  |  |  |
| Number of ADL/IADL limitations | 52 (1.89%) | 2750 | 0 (0.00%) | 1157 | 0 (0.00%) | 5166 | 1 (0.05%) | 2069 |
| Number of ADL limitations | 50 (1.82%) | 2750 | 0 (0.00%) | 1157 | 0 (0.00%) | 5166 | 1 (0.05%) | 2069 |
| Number of IADL limitations | 2 (0.07%) | 2750 | 0 (0.00%) | 1157 | 0 (0.00%) | 5166 | 0 (0.00%) | 2069 |
| No care for ADL/IADL | 1 (0.04%) | 2750 | 23 (1.99%) | 1157 | NA | NA | 1 (0.05%) | 2069 |
| No informal care for ADL/IADL | 11 (0.40%) | 2750 | 47 (4.06%) | 1157 | NA | NA | 1 (0.05%) | 2069 |
| No formal care for ADL/IADL | 21 (0.76%) | 2750 | 48 (4.15%) | 1157 | 7 (0.14%) | 5166 | 1 (0.05%) | 2069 |
| No care for ADL | 3 (0.15%) | 2017 | 0 (0.00%) | 950 | 46 (2.4%) | 1900 | 0 (0.00%) | 1295 |
| No informal care for ADL | 9 (0.45%) | 2017 | 0 (0.00%) | 950 | 44 (2.3%) | 1900 | 0 (0.00%) | 347 |
| No formal care for ADL | 28 (1.39%) | 2017 | 0 (0.00%) | 950 | 4 (0.10%) | 3868 | 0 (0.00%) | 347 |
| No care for IADL | 1 (0.04%) | 2229 | 32 (4.18%) | 765 | NA | NA | 4 (0.23%) | 1716 |
| No informal care for IADL | 14 (0.63%) | 2229 | 50 (6.54%) | 765 | NA | NA | 1 (0.19%) | 520 |
| No formal care for IADL | 14 (0.63%) | 2229 | 51 (6.67%) | 765 | 6 (0.15%) | 3989 | 1 (0.19%) | 520 |

*Notes:* HRS = Health and Retirement Study; ELSA = English Longitudinal Study on Ageing; SHARE = Survey of Health, Ageing and Retirement in Europe (SHARE); CHARLS = China Health and Retirement Longitudinal Study; ADL/IADL= basic or instrumental activities of daily living; ADL = basic activities of daily living; IADL = instrumental activities of daily living.

## Notes

### Competing Interest Statement

The authors have declared no competing interest.

